# Infection and mRNA-1273 vaccine antibodies neutralize SARS-CoV-2 UK variant

**DOI:** 10.1101/2021.02.02.21250799

**Authors:** Venkata Viswanadh Edara, Katharine Floyd, Lilin Lai, Meredith Gardner, William Hudson, Anne Piantadosi, Jesse J. Waggoner, Ahmed Babiker, Rafi Ahmed, Xuping Xie, Kumari Lokugamage, Vineet Menachery, Pei-Yong Shi, Mehul S. Suthar, for the COVID-19 Neutralization Study Group

## Abstract

Antibody responses against the SARS-CoV-2 Spike protein correlate with protection against COVID-19. Serum neutralizing antibodies appear early after symptom onset following SARS-CoV-2 infection and can last for several months. Similarly, the messenger RNA vaccine, mRNA-1273, generates serum neutralizing antibodies that are detected through at least day 119. However, the recent emergence of the B.1.1.7 variant has raised significant concerns about the breadth of these neutralizing antibody responses. In this study, we used a live virus neutralization assay to compare the neutralization potency of sera from infected and vaccinated individuals against a panel of SARS-CoV-2 variants, including SARS-CoV-2 B.1.1.7. We found that both infection- and vaccine-induced antibodies were effective at neutralizing the SARS-CoV-2 B.1.1.7 variant. These findings support the notion that in the context of the UK variant, vaccine-induced immunity can provide protection against COVID-19. As additional SARS-CoV-2 viral variants continue to emerge, it is crucial to monitor their impact on neutralizing antibody responses following infection and vaccination.

---

Antibody responses against the SARS-CoV-2 Spike protein correlate with protection against COVID-19. Serum neutralizing antibodies appear within 10 days post-symptom onset (PSO) following SARS-CoV-2 infection^1^ and are maintained for at least eight months^2^. Similarly, the messenger RNA vaccine, mRNA-1273, generates serum neutralizing antibodies that are detected through at least day 119^3^. The recent emergence of the SARS-CoV-2 B.1.1.7 variant raises concerns about the breadth of these neutralizing antibody responses.

We describe the neutralizing antibody response in 20 acutely infected COVID-19 patients (8-24 days PSO), 20 convalescent COVID-19 individuals (30-90 days PSO), and 14 healthy adult participants that received two injections of the mRNA-1273 vaccine at a dose of 100 µg (18-55 years; day 1 and day 14 post-2nd dose)^4^ against a panel of SARS-CoV-2 variants. This includes an early from Seattle, WA^5^ isolate (USA-WA1/2020), a later D614G variant isolated from a residual nasopharyngeal swab collected from a patient in Atlanta, GA in March 2020 (SARS-CoV-2/human/USA/GA-EHC-083E/2020), and a B.1.1.7 variant isolated from a residual nasopharyngeal swab collected from a patient in San Diego, CA (SARS-CoV-2/human/USA/CA_CDC_5574/2020). A recombinant SARS-CoV-2 containing a single point mutation within the spike at position 501 (Asn to Tyr) was also included^6^. The amino acid differences within the spike protein are described in Figure 1A.

**Figure 1.**
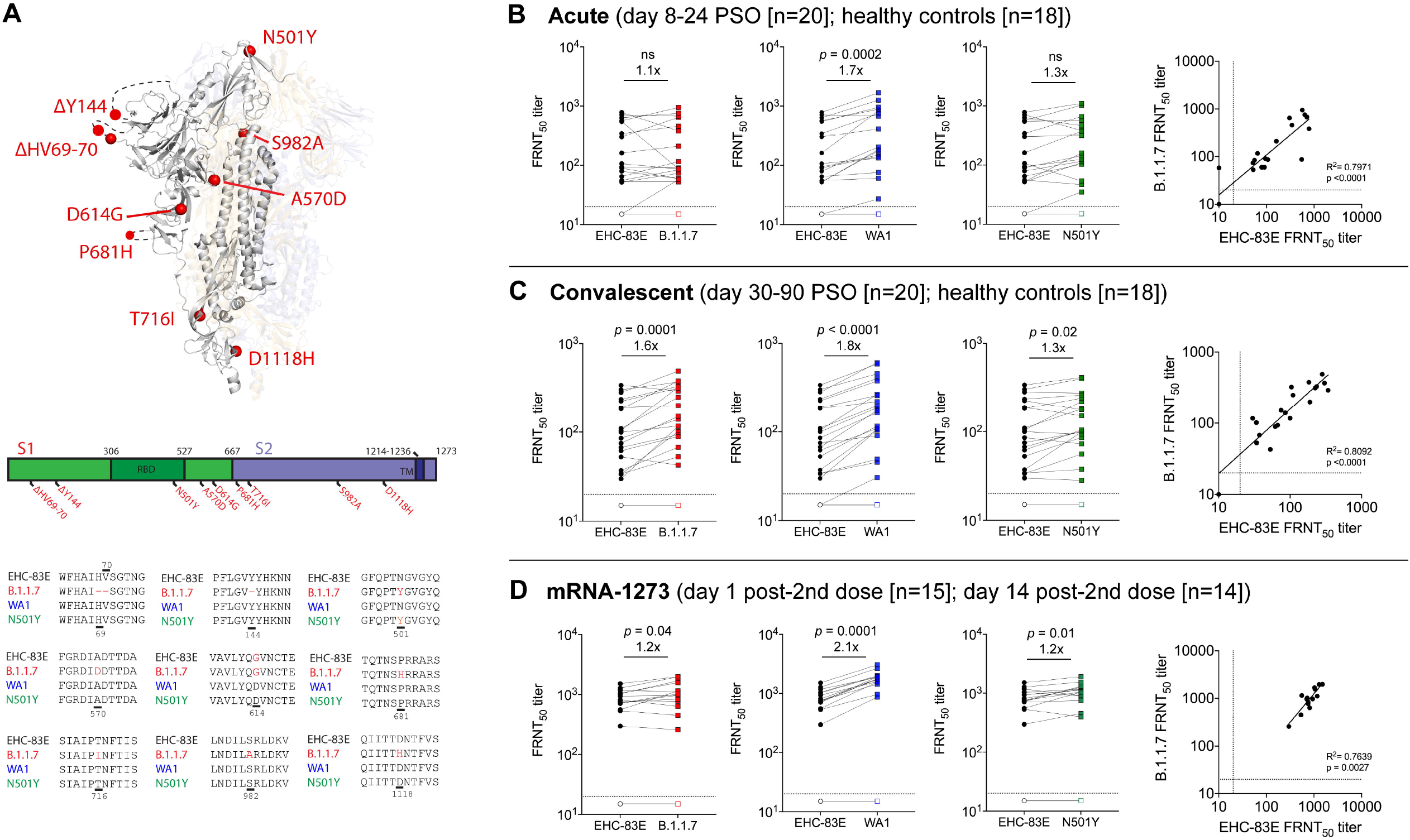
Neutralizing antibody responses against the SARS-CoV-2 B.1.1.7 viral variant. Shown are data from the following cohorts based on natural infection: 20 acutely infected COVID-19 patients (8-24 days PSO; closed symbols), 20 convalescent COVID-19 individuals (30-90 days PSO, closed symbols) 18 healthy controls (open symbols); and individuals that received 100 ug of mRNA-1273 (18 to 55 years old) on day 1 post-2nd dose (15 participants; open symbols) and day 15 post-2nd dose (14 participants; closed symbols). A schematic of the amino acid changes within the spike protein are shown between the SARS-CoV-2 variants (Panel A); The 50% inhibitory titer (FRNT_50_) on the focus reduction neutralization (FRNT) assay for the EHC-83E (black), B.1.1.7 (red), WA1 (blue) and N501Y (green) SARS-CoV-2 variants and correlation plots between EHC-83E and B.1.1.7 viruses are shown for the acutely infected COVID-19 patients (Panel B), convalescent COVID-19 individuals (Panel C), and mRNA-1273 vaccinated individuals (Panel D). Statistical significance was determined using a Wilcoxon paired t-test. The GMT fold change for the respective isolates relative to EHC-83E is shown in each of the plots.

Serum neutralization titers were measured using a live-virus Focus Reduction Neutralization Test (FRNT) assay^7^ and no reduction in the neutralization titers were observed in the variants compared to SARS-CoV-2 EHC-83E (**Figure 1**). The 50% inhibitory dilution (FRNT_50_) geometric mean titer (GMT) was 101, 109, 175 and 133 for EHC-83E, B.1.1.7, WA1, and N501Y, respectively, for acutely infected COVID-19 patients. The FRNT_50_ GMT was 89, 142, 168 and 112 for EHC-83E, B.1.1.7, WA1, and N501Y, respectively, for convalescent COVID-19 individuals. The FRNT_50_ GMT was 804, 965, 1708 and 994 for EHC-83E, B.1.1.7, WA1, and N501Y, respectively, for the mRNA-1273 vaccinated individuals. Neutralization titers correlated between the EHC-83E and B.1.1.7 variants across acutely infected COVID-19 patients (R^2^= 0.7971; p<0.0001), convalescent COVID-19 individuals (R^2^= 0.8092; p<0.0001), and mRNA1273 vaccinated individuals (R^2^= 0.7639; p= 0.0027).

These results show that neutralizing antibody titers following natural infection or vaccination are effective against the UK variant (B.1.1.7) and viral strains containing single point mutations at positions 501 and 614 within the spike protein. These findings support the notion that in the context of the UK variant, vaccine-induced immunity can provide protection against COVID-19. As additional SARS-CoV-2 viral variants continue to emerge, it is crucial to monitor their impact on neutralizing antibody responses following infection and vaccination.

This work was supported in part by grants (NIH P51 OD011132, 3U19AI057266-17S1, U19AI090023, R01AI127799, R01AI148378, K99AI153736, and UM1AI148684 to Emory University; and R00AG049092 and R24AI120942 to University of Texas Medical Branch) from the National Institute of Allergy and Infectious Diseases (NIAID), National Institutes of Health (NIH), by the Emory Executive Vice President for Health Affairs Synergy Fund award, the Pediatric Research Alliance Center for Childhood Infections and Vaccines and Children’s Healthcare of Atlanta, COVID-Catalyst-I^3^ Funds from the Woodruff Health Sciences Center and Emory School of Medicine, Woodruff Health Sciences Center 2020 COVID-19 CURE Award, and the Vital Projects/Proteus funds. The findings and conclusions in this report are those of the author(s) and do not necessarily represent the official position of the Centers for Disease Control and Prevention.

## Data Availability

The datasets generated during and/or analyzed in the current study are available from the corresponding author on reasonable request following acceptance in a peer-reviewed journal.

## Supplementary Appendix

**\*The COVID-19 Neutralization Team Members**

**(listed in pubmed, and ordered alphabetically by institutional affiliation)**

This study was a collective group effort across multiple institutions and locations. Below is a list of sites and staff that significant contributed to sample collection and processing, viral sequencing, design and conduct of the neutralization assays.

<u>Emory University School of Medicine, Atlanta, GA</u>. David S. Stephens, M.D., Evan J. Anderson, M.D., Srilatha Edupuganti, M.D., M.P.H, Nadine Rouphael, M.D., M.Sc., Grace Mantus, M.S., Lindsay Nyhoff, Ph.D., Jens Wrammert, Ph.D., Max W. Adelman, M.D., M.Sc., Rebecca Fineman, B.S., Shivan Patel, M.D., Rebecca Byram, M.E., Dumingu Nipuni Gomes, M.P.H., Garett Michael, B.S., B.A., Hayatu Abdullahi, M.D. Erin M. Scherer, Ph.D., Nour Beydoun, M.D., Bernadine Panganiban, Nina McNair, Kieffer Hellmeister, Jamila Pitts, Joy Winters, Jennifer Kleinhenz, Jacob Usher, James O’Keefe, M.D.

<u>UC San Diego School of Medicine, San Diego, CA.</u> Louise C. Laurent, M.D., Ph.D., Peter De Hoff, Ph.D., Holly Valentine, B.A., M.P.H., Rob Knight, Ph.D., Phoebe Seaver, B.A., M.P.H., Gene W. Yeo, Ph.D., M.B.A., Shashank Sathe, B.Tech., M.S. Aaron Carlin, M.D., Ph.D.

<u>The Scripps Research Institute, La Jolla, CA.</u> Kristian G. Andersen, Ph.D., Mark Zeller, Ph.D., Karthik Gangavarapu, B.S., Catie Anderson, B.A., Alaa Abdel Latif, B.A., M.Phil., B.S.

<u>Centers for Disease Control and Prevention, Atlanta, GA</u>. Natalie Thornburg, Ph.D., Azaibi Tamin, Ph.D., Jennifer L. Harcourt, Ph.D., Maureen Diaz, Ph.D., Suxiang Tong, Ph.D, Ying Tao, Ph.D., Jing Zhang, Ph.D., Phili Wong, M.S., Shilpli Jain, Ph.D., Jennifer Folster, Ph.D.

We thank the mRNA-1273 phase 1 study team and the Division of Microbiology and Immunology for providing clinical samples.

We gratefully acknowledge the following Authors from the Originating laboratories responsible for obtaining the specimen and the Submitting laboratory where genetic sequence data were generated and shared via the GISAID Initiative, on which this research is based.

| **Virus name** | **Accession No** | **Collected** | **Originating laboratory** | **Submitting laboratory** | **Authors** |
| --- | --- | --- | --- | --- | --- |
| hCoV-19/USA/CA-SEARCH-5574/2020 | EPI_ISL_751801 | 2020-12-29 | UCSD<br>EXCITE | Andersen lab<br>at Scripps<br>Research | SEARCH<br>Alliance San<br>Diego |

## Materials and Methods

### Ethics statement

For samples collected at Emory University, collection and processing were performed under approval from the University Institutional Review Board (IRB #00001080 and #00022371). Adults ≥18 years were enrolled who met eligibility criteria for SARS-CoV-2 infection (PCR confirmed by a commercially available assay) and provided informed consent. For the mRNA-1273 phase 1 clinical trial, the neutralization assays were conducted on deidentified specimens, as protocol-defined research. The mRNA-1273 phase 1 clinical trial (NCT04283461) was reviewed and approved by the Advarra institutional review board, which functioned as a single board. The trial was overseen by an independent safety monitoring committee. All participants provided written informed consent before enrollment. The trial was conducted under an Investigational New Drug application submitted to the Food and Drug Administration. The NIAID served as the trial sponsor and made all decisions regarding the study design and implementation.

### Serum samples

For Emory University, acute peripheral blood samples were collected from hospitalized patients at the time of enrollment. Convalescent samples from COVID-19 patients were collected when the patients were able to return for a visit to the clinical research site at the next study visit. Convalescent samples were collected at a range of times (30-90 days) post symptom onset. Serum samples for the mRNA-1273 phase 1 study were obtained from the Division of Microbiology and Infectious Diseases, National Institute of Allergy and Infectious diseases for the mRNA-1273 phase 1 study team and Moderna Inc. Study protocols and results were previously reported^8^. Samples tested were collected from 15 healthy individuals on day 1 and day 14 post-2^nd^ dose of the mRNA-1273 vaccine.

### Viruses and cells

VeroE6 cells were obtained from ATCC (clone E6, ATCC, #CRL-1586) and cultured in complete DMEM medium consisting of 1x DMEM (VWR, #45000-304), 10% FBS, 25mM HEPES Buffer (Corning Cellgro), 2mM L-glutamine, 1mM sodium pyruvate, 1x Non-essential Amino Acids, and 1x antibiotics. The infectious clone SARS-CoV-2 (icSARS-CoV-2), derived from the 2019-nCoV/USA_WA1/2020 strain, was propagated in VeroE6 cells (ATCC) and sequenced^9^. The B.1.1.7 variant (SARS-CoV-2/human/USA/CA_CDC_5574/2020) was isolated from a residual nasopharyngeal swab collected from a patient in San Diego, CA, propagated in Vero cells, sequenced and kindly provided by Natalie Thornburg (CDC, Atlanta, GA). EHC-083E was isolated from a residual nasopharyngeal swab collected from a patient in Atlanta, GA in March 2020 (SARS-CoV-2/human/USA/GA-EHC-083E/2020), plaque purified on Vero cells, propagated on Vero cells and sequenced. The sequences of the working viral stocks were identical to the sequence obtained in the nasopharyngeal swab. The N501Y SARS-CoV-2 virus, a recombinant SARS-CoV-2 containing a single point mutation within the spike at position 501 (Asn to Tyr), was kindly provided by Dr. Pei-Yong Shi^6^. Viral titers were determined by focus-forming assay on VeroE6 cells. Viral stocks were stored at -80°C until use.

### Focus Reduction Neutralization Assay

FRNT assays were performed as previously described^7^. Briefly, samples were diluted at 3-fold in 8 serial dilutions using DMEM (VWR, #45000-304) in duplicates with an initial dilution of 1:10 in a total volume of 60 μl. Serially diluted samples were incubated with an equal volume of SARS-CoV-2 (100-200 foci per well) at 37° C for 1 hour in a round-bottomed 96-well culture plate. The antibody-virus mixture was then added to Vero cells and incubated at 37° C for 1 hour. Post-incubation, the antibody-virus mixture was removed and 100 µl of prewarmed 0.85% methylcellulose (Sigma-Aldrich, #M0512-250G) overlay was added to each well. Plates were incubated at 37° C for 24 hours. After 24 hours, methylcellulose overlay was removed, and cells were washed three times with PBS. Cells were then fixed with 2% paraformaldehyde in PBS (Electron Microscopy Sciences) for 30 minutes. Following fixation, plates were washed twice with PBS and 100 µl of permeabilization buffer (0.1% BSA [VWR, #0332], Saponin [Sigma, 47036-250G-F] in PBS), was added to the fixed Vero cells for 20 minutes. Cells were incubated with an anti-SARS-CoV spike primary antibody directly conjugated to biotin (CR3022-biotin) for 1 hour at room temperature. Next, the cells were washed three times in PBS and avidin-HRP was added for 1 hour at room temperature followed by three washes in PBS. Foci were visualized using TrueBlue HRP substrate (KPL, # 5510-0050) and imaged on an ELISPOT reader (CTL).

### Quantification and Statistical Analysis

Antibody neutralization was quantified by counting the number of foci for each sample using the Viridot program^10^. The neutralization titers were calculated as follows: 1 - (ratio of the mean number of foci in the presence of sera and foci at the highest dilution of respective sera sample). Each specimen was tested in duplicate. The FRNT-50 titers were interpolated using a 4-parameter nonlinear regression in GraphPad Prism 8.4.3. Samples that do not neutralize at the limit of detection at 50% are plotted at 10 and was used for geometric mean calculations.

